# Four Years of Continuous Weekly Wastewater Analysis Reveals Sequencing as a Robust Alternative to PCR for Respiratory Virome Surveillance

**DOI:** 10.64898/2026.10.13.26364774

**Authors:** Harihara Prakash, Vasanthi Avadhanula, Michael J. Tisza, Juwan Cormier, Matthew C. Ross, Ryan K. Perez, James D. Chang, Camille R. Mazurek, Jennifer Deegan, Joe F. Petrosino, Pedro A. Piedra, Justin R. Clark, Eric Boerwinkle, Anthony W. Maresso

## Abstract

Wastewater-based epidemiology uses both targeted and non-targeted bioanalytical approaches to detect and track disease-causing microbes in wastewater. Targeted approaches such as quantitative polymerase chain reaction (qPCR) remain widely used while sequencing-based approaches have slowly gained popularity. However, concordance between established targeted molecular assays and sequencing approaches remains incompletely characterized. To assess the validity of sequencing as an alternative to qPCR, we compared outcomes of both methods targeting five respiratory viruses in wastewater samples collected across 16 Texas cities over 4 years (May 2022-July 2026) of continuous sampling. We found positive correlations between qPCR and sequencing measurements, with both methods independently correlating with statewide clinical emergency department visits. qPCR positivity also increased with genomic sequencing coverage, and sequencing demonstrated greater potential than qPCR as an early respiratory virus outbreak detection method. Our findings support metagenomic sequencing as a sensitive and highly specific alternative to targeted qPCR for wastewater surveillance, and reinforce its utility towards long-term longitudinal monitoring of respiratory viruses.

## Introduction

Modern-day microbial metagenomics employs a vast array of bioanalytical techniques to assess microbial species composition in environmental samples^1^. Using both targeted and non-targeted detection approaches, researchers today analyze samples collected from the environment, looking for specific clinically relevant microbes of interest^2^. Targeted approaches such as quantitative PCR (qPCR) and droplet digital PCR (ddPCR) use target-specific primer sequences to quantify microbial abundance, and remain the gold standard for detection of genomic material^3,4^. These approaches enable both detection of relevant microbes and identification of critical virulence-associated genetic elements, including antimicrobial resistance genes and virulence factors^5,6^.

While targeted approaches are often used, non-targeted approaches have slowly gained popularity in wastewater metagenomics over the last 15 years^7^. With the rapid development of modern-day sequencing technologies, the ability to achieve improved sequencing depth and breadth of microbial genomes has increased dramatically^8^. Consequently, this allows the detection of multiple organisms in a sample using sequencing methods rather than being constrained to a single species with targeted PCR methods. Probe panel-based hybrid-capture sequencing, for instance, can be used to detect a wide array of microbial targets within the same sample (3,153 viral targets with TWIST Bioscience’s Comprehensive Viral Research Panel^9^). Its broad target range makes it a cost- and time-effective technique suitable for simultaneous surveillance of multiple clinically relevant microbes in wastewater^10^.

Sewage had long been known to carry a readable record of community infection^11^, but wastewater surveillance moved to the center of public health practice during the COVID-19 pandemic, when SARS-CoV-2 RNA proved detectable^12,13^, quantifiable across an entire metropolis as a program^14,15^, and ultimately sequenceable at the level of variants and whole viromes^10,16^. As SARS-CoV-2 spread from early 2020, wastewater monitoring scaled from individual studies into sustained local^15,17,18^ and national^19,20^ municipal surveillance programs.

Sequencing those same samples resolved variants circulating in each community and in some cases detected cryptic variant transmission before clinical genomic surveillance^10,15,16,21,22^. Today, studies conducted across the Americas, Asia, Europe and Oceania have highlighted the success of SARS-CoV-2-targeted wastewater-based epidemiology (WBE)^12–14,23,24^. Building off the success of this viral monitoring system, the list of target viruses suitable for surveillance expanded, and tracking mpox, avian influenza, respiratory syncytial virus (RSV) and more was initiated worldwide^25–28^.

In May 2022, the Texas Epidemic Public Health Institute (TEPHI) funded the Texas Wastewater Environmental Biomonitoring (TexWEB) initiative^29^, the first and longest running continuous wastewater metagenomics program in the U.S. The goals of this program were to leverage sequencing-based approaches to monitor viral prevalence in wastewater and provide an early warning system for outbreaks for the entire human virome, periodically notifying health officials of concerning viral trends^30^. As of July 2026, TexWEB researchers successfully detected and tracked hundreds of viruses at once, including but not limited to human immunodeficiency virus (HIV), avian influenza, mpox, measles, parvovirus B19, and cancer-causing viruses, and have also tracked non-endemic viruses during high-travel periods such as during the 2026 FIFA World Cup^10,31,26,32–35^. By updating interactive dashboards displaying statewide viral trends weekly, TexWEB provides publicly accessible tools for monitoring changes in viral abundance over time^36,37^. The program’s sustained longitudinal sampling and all-virus detection approach has also enabled the discovery of new seasonal patterns within the collective virome^38^.

Sequencing-based methods show great promise to be an ideal “radar” system for pathogen activity. However, such methods must stand up to the gold standard that dominates public health efforts. Whereas prior work has compared qPCR against various sequencing approaches^2,39,40^ and has aligned method-specific detections with clinical incidence^41,42^, these few studies only cover a small collection of viruses in modest sample sets obtained over less than a year. To build a comparative framework between these methods, we prospectively tested wastewater from 40 sites across 16 Texas cities over four years with both targeted qPCR and hybrid-capture sequencing for five respiratory viruses (influenza A, influenza B, RSV A, RSV B and SARS-CoV-2)^43^. Our findings suggest that across four years of surveillance, hybrid-capture sequencing detected target respiratory viruses on average earlier than qPCR, showing promise as an early warning system compared to traditional approaches. Sequencing also recovered viral genetic material more consistently year-round and correlated closely with both qPCR measurements and clinical indicators, highlighting its ability to complement or potentially replace PCR-based approaches for wastewater based viral surveillance.

## Results

### Collection, analysis and curation of nearly 5,000 wastewater samples

Starting in May 2022, wastewater samples were collected weekly at wastewater treatment plants across the state of Texas, USA. As of July 2026, TexWEB facilitates consistent sampling of 16 Texas cities **(Fig. 1A)**. Houston, El Paso and Lubbock were among the first cities where sampling was initiated in 2022. From these three cities alone, over 3,000 samples have been collected to date. As the program grew over 4 years, more cities joined the initiative, contributing to a current total of 4,935 samples collected over 40 sites. Around 4-7 samples were collected per site monthly, representing weekly sampling and occasional cases when more than one sample was collected from a site **(Fig. 1B)**.

**Figure 1:**
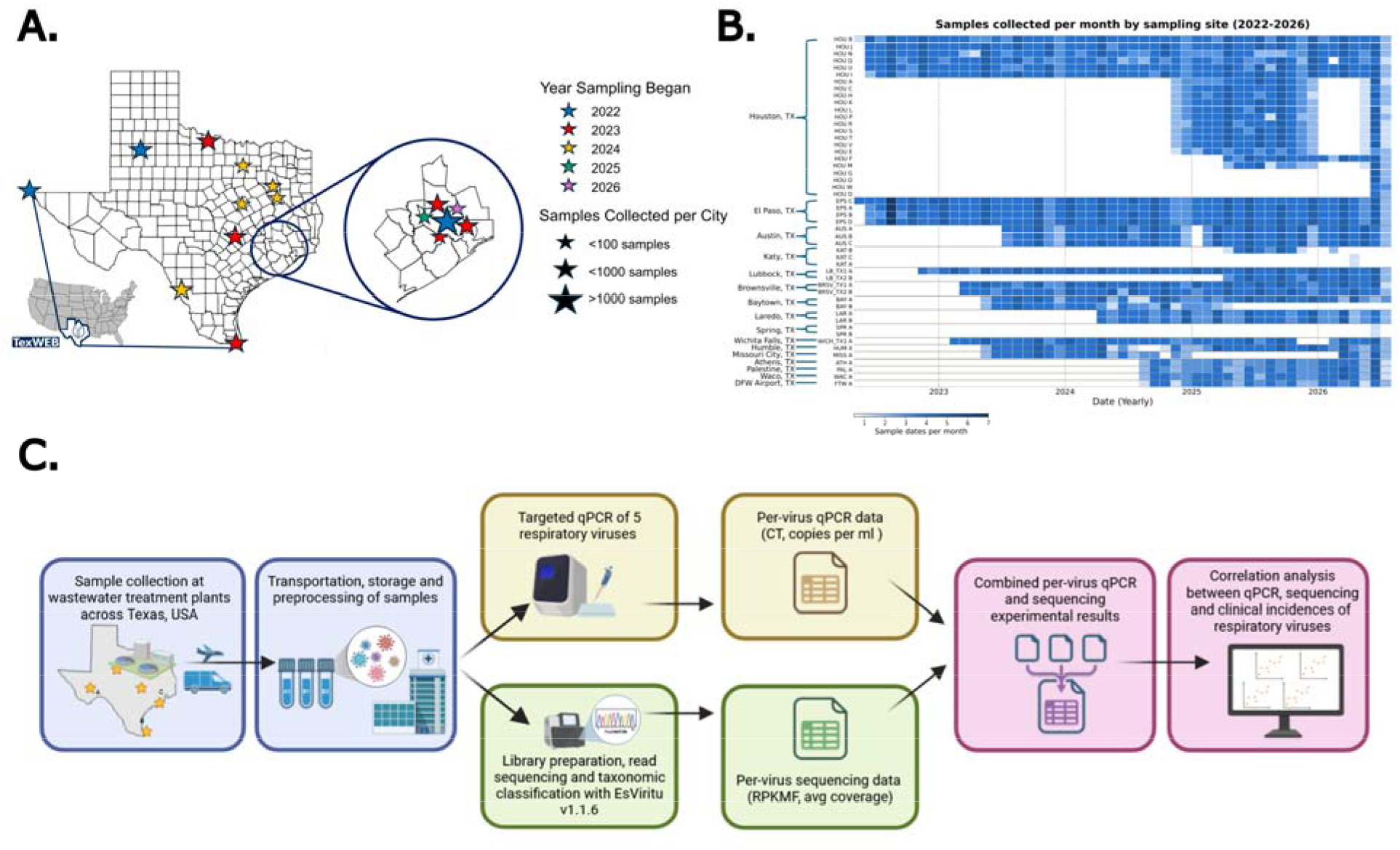
Overview of TexWEB wastewater sample collection network and study methodology. **(A)** represents the Texas county map with sampling cities as of July 2026. Stars represent cities where wastewater samples are collected. Star colors are coded by the year the first sample was collected from any site at that city. Star size is proportional to the number of samples collected from that city. **(B)** represents samples collected per month by sampling site from 2022-2026. Number of samples collected each month per site starting from 2022 up to 2026 are represented by a single cell. Sites are ordered b city with the largest number of sites to the lowest number of sites. Darker shades represent more sample collected from that site at that month, while no color represents no samples collected at that site for that month. **(C)** represents the project methodology from sampling to per-virus data analysis. Blue boxes represent the sample collection and transport. Green boxes highlight the sequencing pipeline and its output units of measurement, while yellow boxes represent the qPCR process with its corresponding outputs. Finally, the pink boxes show the joining of the two results and subsequent correlation analysis. See Methods for in-depth section-specific information.

All collected samples were transported to Baylor College of Medicine in Houston, Texas for qPCR and hybrid-capture sequencing **(Fig. 1C)**. Once tested, a virus may be either detected or not detected in the sample. Of the 4,935 samples collected as of July 2026, nearly 4,000 were tested with both sequencing and qPCR for influenza A, influenza B, RSV A, RSV B and SARS-CoV-2. Around ∼900 samples were tested with either sequencing or qPCR, but not both, and were excluded from the study. After metadata correction, removal of artifacts, and overall data curation, a total of 3910, 3897, 3879, 3914, and 3910 samples for the influenza A, influenza B, RSV A, RSV B and SARS-CoV-2 respectively were used for correlation analyses. These samples thus represent a corpus of data processed by both techniques from which direct comparisons can be carried out.

### Variations in detection capability between qPCR and sequencing

Across all 5 respiratory viruses, differences in detection capabilities were present among the samples treated with both qPCR and the sequencing **(Fig. 2)**. Interestingly, influenza A, influenza B, RSV A and RSV B were not detected in most samples tested by both methods (62-81% of all samples per virus). The same could not be said for SARS-CoV-2, where a substantial 78% of wastewater samples tested with both techniques came back positive, a trend unseen across the other respiratory viruses. Only 5% of the samples tested for SARS-CoV-2 tested negative for the virus with both techniques. This likely reflects the relatively high abundance and year-round persistence of SARS-CoV-2 in wastewater.

**Figure 2:**
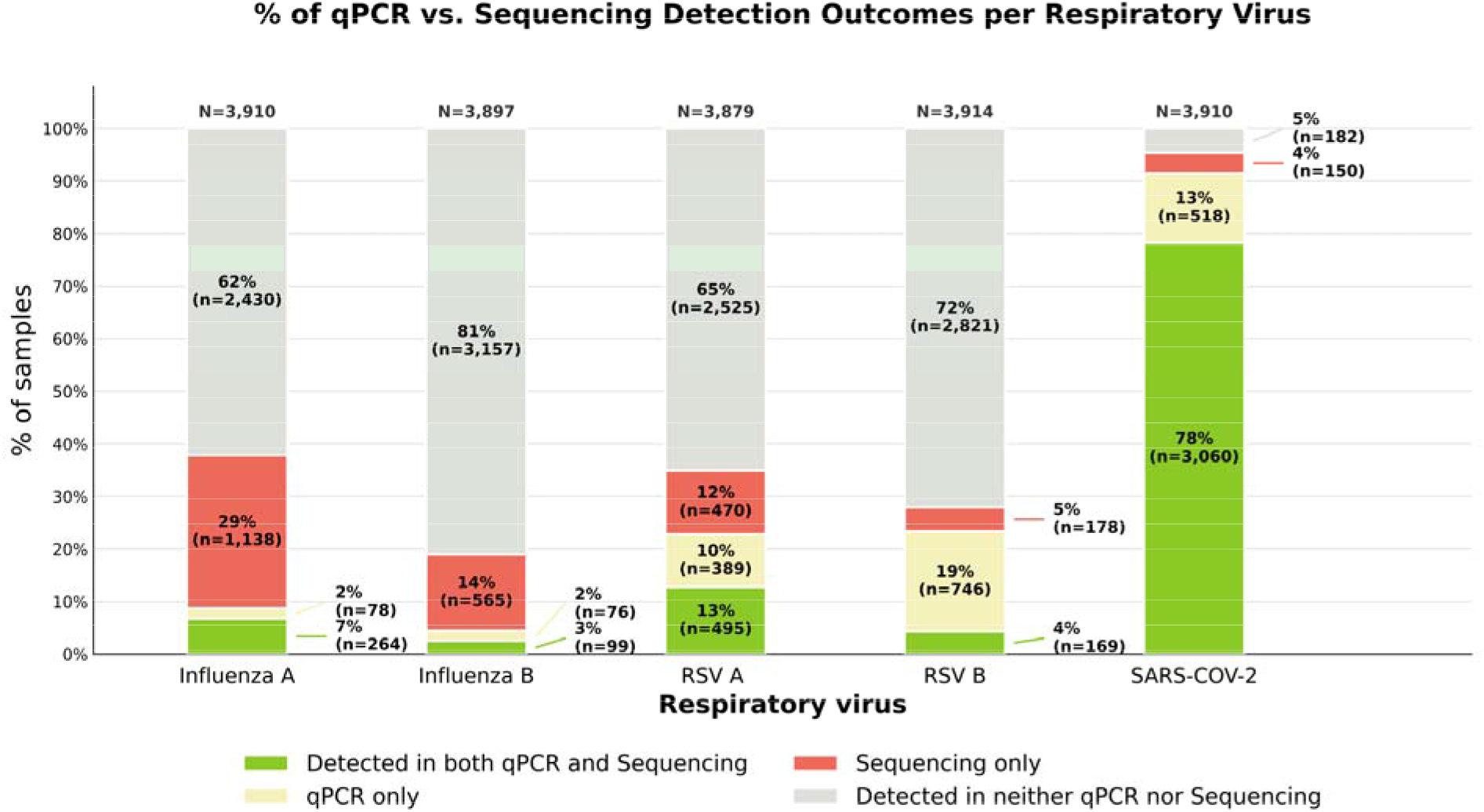
Number of virus-positive samples when tested with qPCR and sequencing per respiratory virus. Bar chart shows the number of samples that tested positive for target viruses by each method. Each bar represents the % of samples tested for each respiratory virus using qPCR and sequencing. Each stac within a full bar represents whether the corresponding respiratory virus was detecting the sample with qPCR only, sequencing only, both techniques and by neither technique. A slight mismatch in the number of samples tested between viruses arises from minor differences in assay completion.

For the influenza viruses, we observed that sequencing detected target viruses in more samples relative to qPCR. For influenza A, sequencing-only detections were nearly 30% of all samples, while qPCR-only detections were only 2%. Similarly, influenza B’s sequencing-only detections were 14% of all samples tested, and qPCR-only detections were also only 2% of the samples. In addition, samples where both methods detected the virus were few, and mostly, neither method detected the virus (possibly due to seasonality of respiratory viruses since we did not exclude “off-season” samples from our analysis; see Discussion). The results for RSV, however, were different. For RSV A, 13% of samples were detected by both methods, 10% by qPCR alone and 12% by sequencing alone. In contrast, RSV B qPCR detections were higher than sequencing detections, 19% vs. 5%, thereby demonstrating the ability of qPCR to better detect RSV B over sequencing. We see that across all 5 viruses, variations in detection ability exist between methods, with RSV more likely to be detected by qPCR while influenza is more likely to be detected by sequencing.

### Strong concordance of qPCR and sequencing across multiple viruses

To visualize the correlation between qPCR and sequencing results, we built scatter plots per target respiratory virus. Here, we used Spearman’s rank correlation to quantify correlation due to the non-normal nature of qPCR and sequencing data as shown by Shapiro-Wilk’s test **(Shapiro’s W = 0.01-0.58, p < 0.001; Supp. Table 1)**.

First, we plotted all recorded data points regardless of detection results **(Fig. 3)**. When correlating all samples across all viruses, we observed a moderate positive correlation (Spearman’s ρ = 0.560, p < 0.001) **(Fig. 3A)**. Next, we plotted per-virus scatter plots, assessing the correlation between qPCR and sequencing for each **(Fig. 3B-F)**. We found the strongest correlation in SARS-CoV-2 (Spearman’s ρ = 0.702, p < 0.001), followed by RSV A (Spearman’s ρ = 0.443, p < 0.001), influenza A (Spearman’s ρ = 0.303, p < 0.001), influenza B (Spearman’s ρ = 0.240, p < 0.001) and RSV B (Spearman’s ρ = 0.194, p < 0.001).

**Figure 3:**
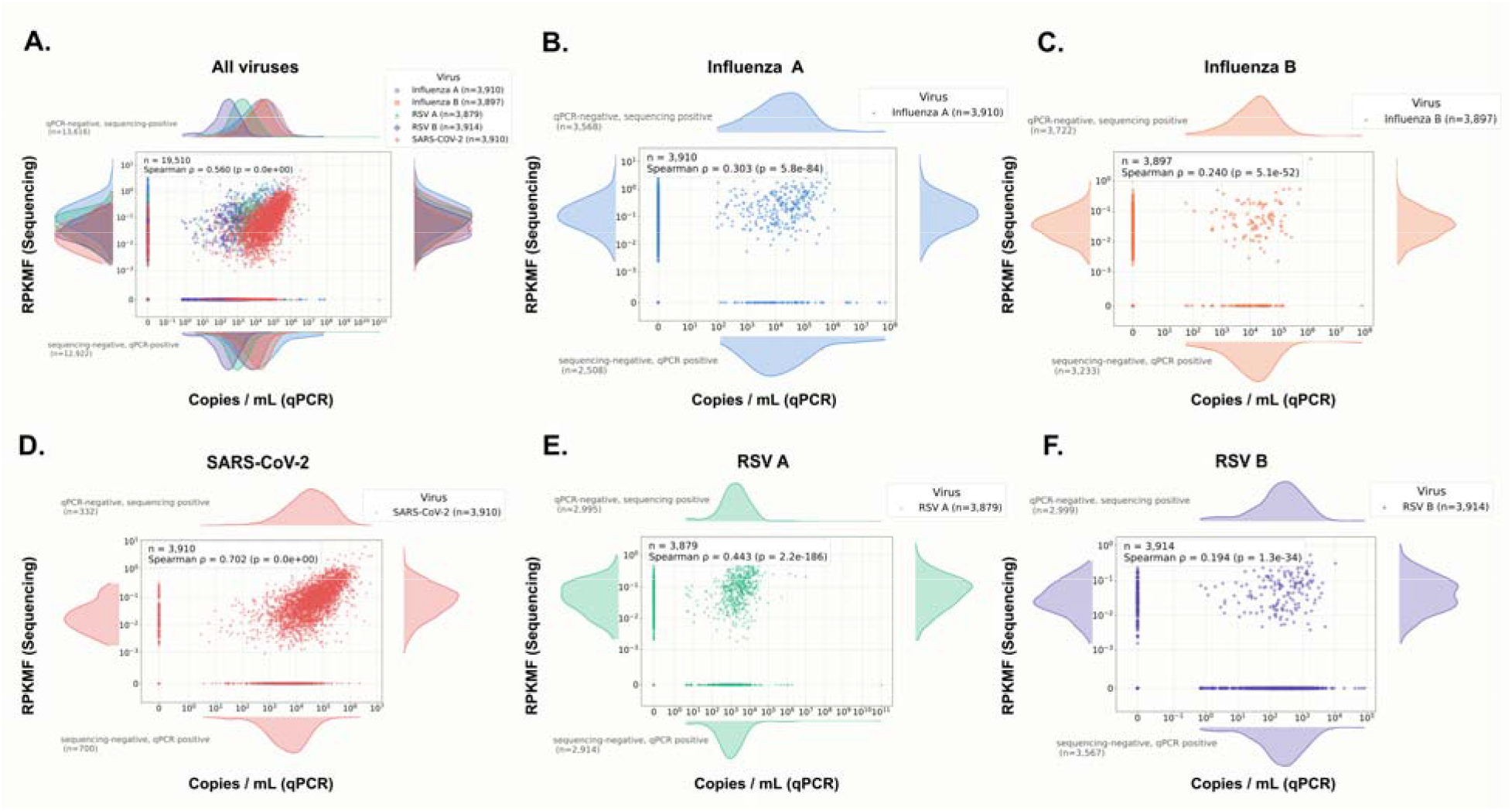
Correlation of qPCR vs. Sequencing results across all respiratory viruses. Scatter plots plotted on a symlog axis to accommodate zero values denote the correlation of all sample tested with both qPCR and sequencing for 5 respiratory viruses. Axis-oriented violin plots highlight the data distribution. Top and Right violin plots represent the distribution of nonzero qPCR and sequencing outcomes respectively. Left and Bottom violin plots represent distribution of qPCR-negative/sequencing-positive and sequencing-negative/qPCR-positive data points respectively. **(A)** denotes the correlation between qPCR and sequencing of all sample datapoints for all 5 viruses. **(B-F)** represent per-virus correlation for all data points. These include cases where neither method detected the virus, when either method detected the virus, and when both methods detected the virus.

To assess the correlation between those samples where both qPCR and sequencing tested positive for the virus, we looked at positive-only samples **(Supp. Fig 1)**. On plotting scatter plots for positive-only samples for all viruses taken together, we found a moderate positive correlation between qPCR and sequencing (Spearman’s ρ = 0.465, p < 0.001; **Supp. Fig 1A**). When assessing per-virus correlation between qPCR and sequencing, we found a similar trend as Fig. 3 with a minor variation **(Supp. Fig. 1B-F)**. Here, SARS-CoV-2 had the strongest correlation (Spearman’s ρ = 0.645, p < 0.001), followed by RSV A (Spearman’s ρ = 0.393, p < 0.001), influenza A (Spearman’s ρ = 0.273, p < 0.001), RSV B (Spearman’s ρ = 0.209, p < 0.01) and finally influenza B (Spearman’s ρ = 0.105, p = 0.3). Together, this data suggests that sequencing-based detections of respiratory viruses correlate well with qPCR detections, the current gold-standard assay in the field.

### Strong concordance of clinical cases with sequencing detections

To assess which method more accurately reflected statewide respiratory virus trends, we independently correlated qPCR and sequencing measurements with annual statewide clinical case data from Texas. Sourced from the Texas Department of State Health Services (DSHS), we used emergency department visit rates (clinical ED rates) for influenza, RSV and SARS-CoV-2 as our clinical case metric^44^. As clinical data was only available for influenza and RSV taken together, we combined our sequencing and qPCR detections of influenza A and influenza B as well as RSV A and RSV B respectively for our analysis.

First, we found that taken weekly, sequencing outcomes aligned extremely well with % of clinical ED rates across all three viruses, with nearly every major clinical peak overlapping with a wastewater peak from sequencing data **(Fig 4A-C)**. The strongest correlation observed was with SARS-CoV-2 (Spearman’s ρ = 0.91, p < 0.001), followed by influenza (Spearman’s ρ = 0.70, p < 0.001), and RSV (Spearman’s ρ = 0.67, p < 0.001).

**Figure 4:**
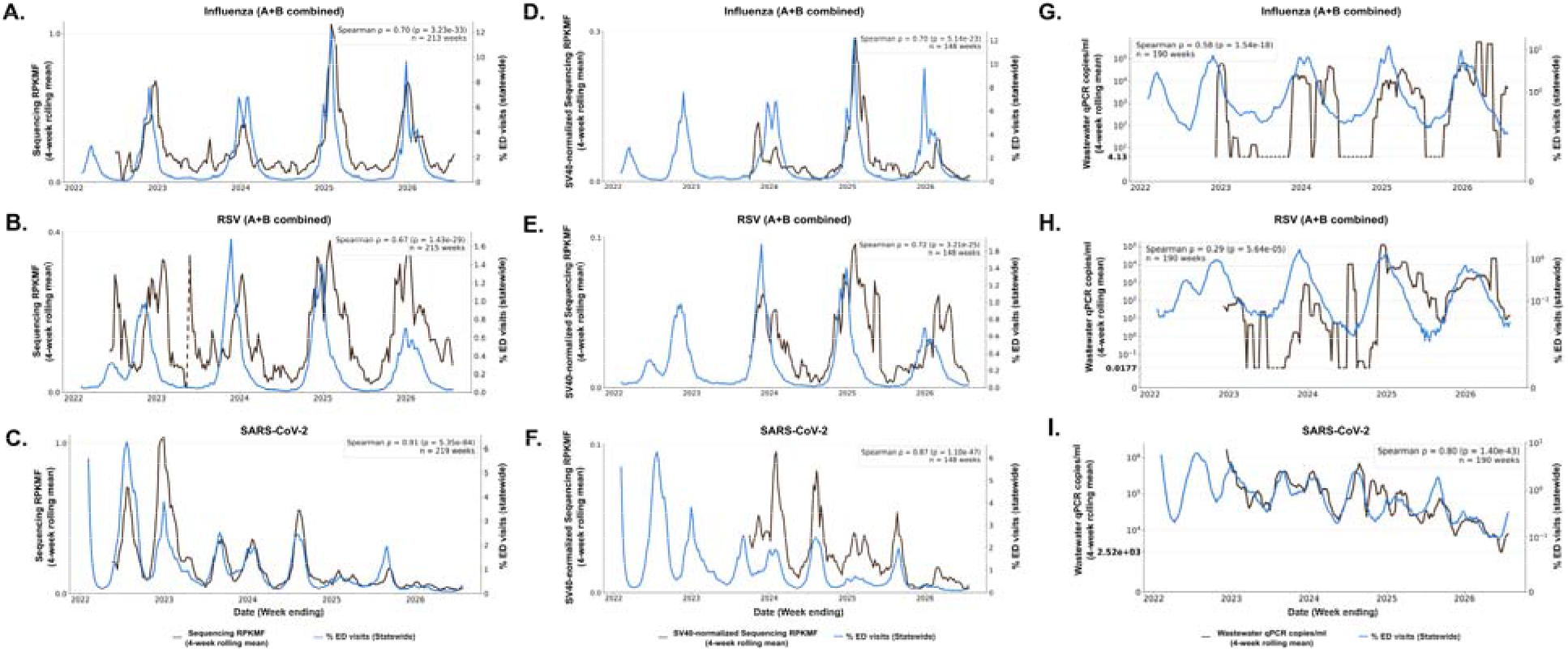
Correlation of sequencing and qPCR with statewide clinical cases. Time-series plots represent the rolling 4-week average abundance of **(A-C)** sequencing vs. clinical ED visit rates, **(D-F)** SV40-normalized sequencing vs. clinical ED visit rates, **(G-I)** and wastewater qPCR vs. clinical ED visit rates. To align with the grouped clinical RSV and influenza data, qPCR- and sequencing-specific RSV and influenza data were also merged (see Methods). Number of weeks is represented b ‘n’, and Spearman’s correlation coefficient was computed across 4-week rolling averages where both variables were non-zero. Dotted lines represent weeks when corresponding viral detection with respective methods was negative.

Since RPKMF is a relative abundance metric, quantified viral abundance exists as a measurement relative to all other viruses in the sample (see Methods). To correct for this, SV40 (Simian Virus 40) spike-in controls were added to samples prior to sequencing from September 2023 onwards. To get the absolute abundance of the target virus per sample, we calculated the SV40-normalized viral abundance, correlating these with clinical ED rates **(Fig 4D-F)**. We found the change in correlation from relative to absolute to be small, with SARS-CoV-2 remaining the strongest correlation (Spearman’s ρ = 0.87, p < 0.001), followed by RSV (Spearman’s ρ = 0.72, p < 0.001) and influenza (Spearman’s ρ = 0.7, p < 0.001).

Finally, when testing the correlation between quantified qPCR outcomes and clinical ED rates, we found that SARS-CoV-2 detected by qPCR correlated strongly with clinical ED rates (**Fig. 4G**, Spearman’s ρ = 0.8, p < 0.001). Interestingly, while influenza still had a moderate positive correlation (**Fig. 4H**, Spearman’s ρ = 0.58, p < 0.001), RSV had the poorest correlation with clinical ED rates (**Fig. 4I**, Spearman’s ρ = 0.29, p < 0.001).

In addition to independently correlating sequencing and qPCR with clinical ED rates, we also looked at the correlation between clinical ED rates and qPCR/sequencing for those samples where both methods had been tested (**Supp. Fig 2, Supp. Table 2**). Here, we found that sequencing correlated better with clinical ED visits than qPCR across all three viruses. Taken together, the strong correlation of sequencing detections with clinical data provides orthogonal validation that the sequencing-based approach approximates the clinical burden for these respiratory viruses.

### Detection of virus signals may relate to the survivability of nucleic acid across the viral genome

qPCR detection in wastewater depends on survival of single short target regions, whereas hybrid-capture sequencing uses tiled probes across the entire genome. This increases the probability of detecting any random genomic fragments that survive the stages of wastewater processing and assay workup. Better sequencing coverage would increase the likelihood that PCR primers land on genomic fragments that survived wastewater and/or its processing. To assess the relationship between genomic coverage (genomic span) and qPCR detection, we compared the qPCR positivity rates with sequencing-derived genomic span **(Fig. 5)**. Genomic span values for all sequencing-positive samples ranged between 0 to 1 and represent the fraction of the viral genome covered by sequencing.

**Figure 5:**
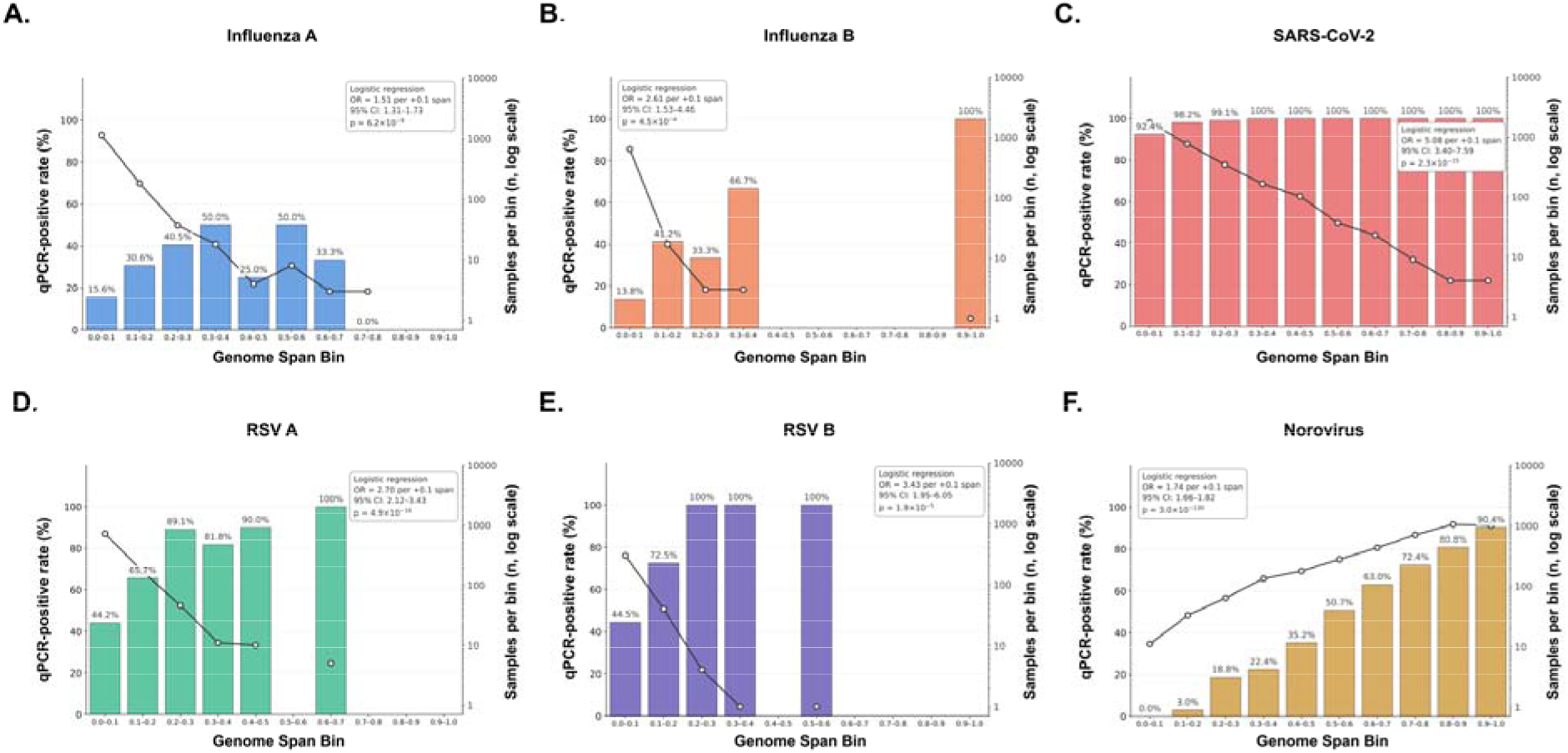
Assessment of the relationship between qPCR positivity rate of target virus genomes with genomic span (breadth) in wastewater. **(A-F)** represent bar charts showing the distribution of qPCR positive-samples among sequencing-detected samples per respiratory virus. The genomic span of each virus per sample was calculated using Lander-Waterman assumptions (See Methods) and represents the coverage of the sequenced read across the full reference genome used for taxonomic classification. Total number of sequencing-positive samples within the genomic bin range is denoted by the line graphs on the secondary y axis, and the qPCR-positive rate above each bar represents the percentage of total sequencing-positive samples that were also qPCR-positive.

We found that across all viruses, qPCR positivity rate increased with genome span **(Supp. Fig. 3)**. On plotting bar charts per target virus, we found that as genome span increased (i.e. the higher the genome positions covered per viral genome), the number of qPCR-positive samples relative to the sequencing-positive samples also increased **(Fig. 5A-E)**. For influenza A, positivity rose from 15.6% in the 0–0.1 bin to 50.0% at 0.3– 0.4; for influenza B, from 13.8% to 66.7%; for RSV A, from 44.2% to 90.0% at 0.4–0.5; and for RSV B, from 44.5% to 100% by 0.2–0.3.

For SARS-CoV-2, qPCR detections were consistently high throughout all genomic span bins **(Fig 5C**), with a comparatively less dramatic increase in positivity rate with increase in genomic span. This contrasted with the other respiratory viruses, where a much more dramatic change in qPCR detection rates was observed with increasing genomic span. To see if SARS-CoV-2 detection trends were influenced by its year-round presence in wastewater, we looked at Norovirus, another enteric virus frequently present in wastewater in high abundances through fecal shedding^45^. Unlike SARS-CoV-2, norovirus exhibited a more pronounced increase in qPCR positivity with increasing genomic span, highlighting the relationship between the completeness of viral reads circulating in wastewater and qPCR detection ability **(Fig. 5F)**. However, like SARS-CoV-2, norovirus showed no seasonality in a coarse Texas-wide composite, unlike the respiratory viruses, suggesting low-level circulation and/or increase detectability between outbreaks **(Supp. Fig. 4)**.

To statistically assess the observed trend of qPCR positivity rate increasing with genomic span, we fit a logistic regression model and computed the odds ratio (OR) for each virus along with 95% confidence intervals. We found that across all five viruses, each 0.1 (10%) increase in span was associated with higher odds of qPCR detection: influenza A OR 1.51 (95% CI 1.31-1.73), influenza B 2.61 (1.53-4.46), RSV A 2.70 (2.12-3.43), RSV B 3.43 (1.95-6.05) and SARS-CoV-2 5.08 (3.40-7.59) where all p < 0.001 **(Fig. 5A-E; Supp. Table 4, Sheet 1)**. The highest odds ratio was observed in SARS-CoV-2, reflecting its near-complete qPCR positivity across all genome span bins. To account for non-independent sample collection, we reran the analysis calculating standard errors when sites were clustered and found that all associations remained significant (all p < 0.03; **Supp. Table 4, Sheet 2**).

In summary, this data suggests that the ability of PCR to detect its cognate viral target is largely dependent on whether that specific genomic segment has survived the transit through wastewater system and downstream processing. The greater the proportion of the genome recovered by sequencing, the better the PCR detection rate. This supports the use of hybrid-capture probe techniques for low abundance signals because hundreds to thousands of probes are tiled across the genome and detection is not dependent on survival of one region, and fragments from any part of the genome can contribute to detection.

### Sequencing detects target respiratory viruses earlier than qPCR

Finally, we determined which of the two methods consistently produced the first positive detection across all samples during the start of the respiratory illness season for each target respiratory virus. This can inform public health action by providing a sufficient lead time during the flu season, when clinical enrollment for these viruses increases. According to the CDC, the respiratory illness season begins around October when cases begin to rise, peaking during the winter months of December and January, and subsequently declining by April^46^. Hence, for each respiratory illness season across 4 years of sampling, we looked at whether qPCR or sequencing detected the target respiratory virus first.

We found that sequencing detected target viruses consistently earlier than qPCR at the beginning of the respiratory illness season. For influenza A, sequencing enabled detection of the virus 35 days before qPCR detection during the 2023-2024 season, and 47 days during the 2024-2025 season **(Fig. 6A)**. Interestingly, during the latest 2025-2026 season, both techniques detected the virus during the start of the flu season simultaneously. A similar trend was observed for influenza B and RSV A **(Fig. 6B-C)**. For RSV B however, simultaneous detection occurred in 2023-2024 season followed by a lead of 15 and 6 days where sequencing enabled earlier detection **(Fig.6D)**. While a delay in detecting these viruses was present among the two methods, we found that the monthly detection rates of qPCR and sequencing correlated reasonably well with each other across all viruses (Spearman’s ρ = 0.52-0.74, all p < 0.001).

**Figure 6:**
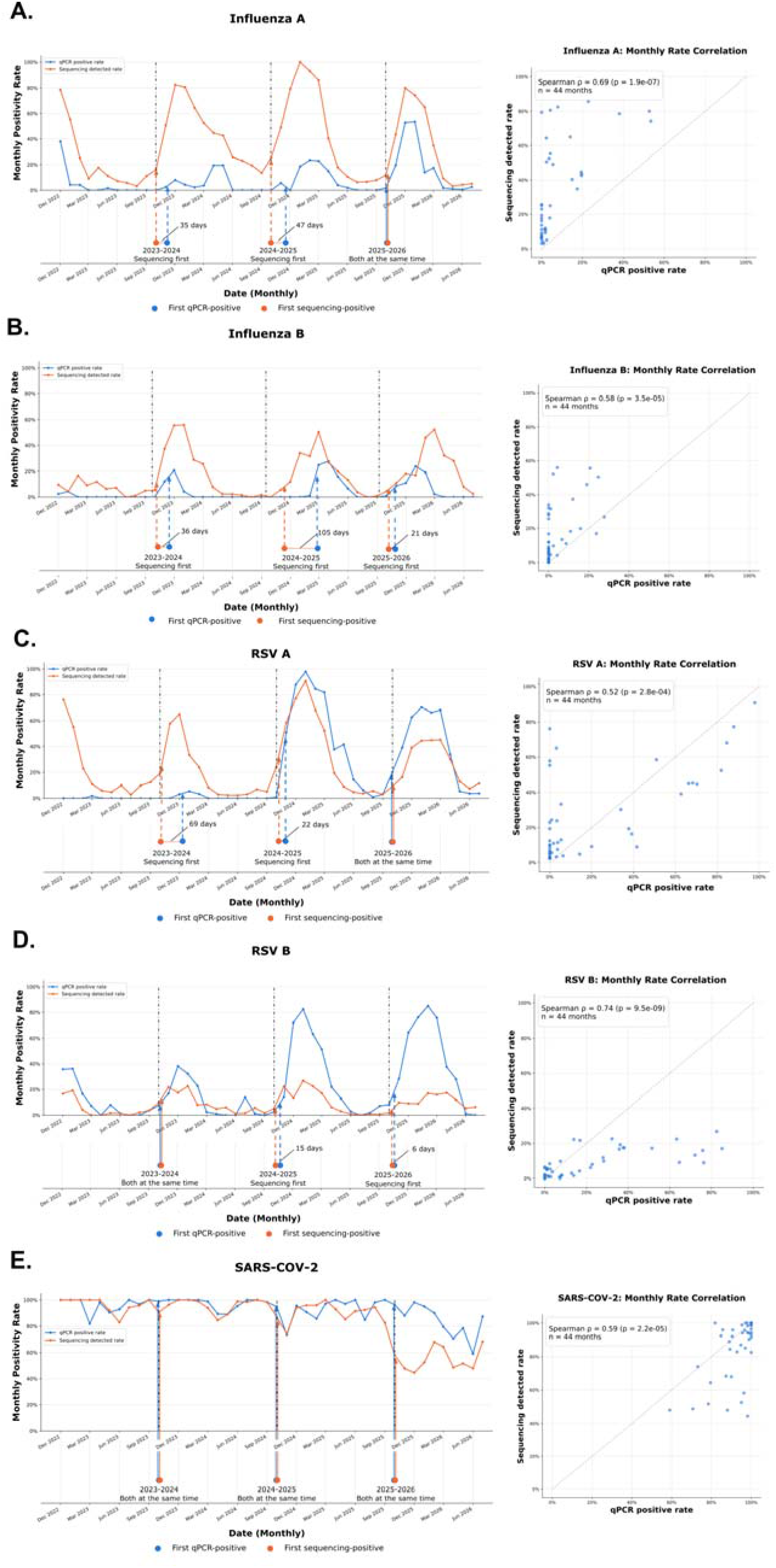
Comparing virus detection timing between sequencing and qPCR detection methods. Time-series charts represent the monthly positivity rate of samples for **(A)** influenza A; **(B)** influenza B; **(C)** RSV A; **(D)** RSV B; and **(E)** SARS-CoV-2 dynamics respectively using sequencing (orange) and qPCR (blue). Dumbbell plots aligned beneath the time-series plot represent the time point at when each method first detected the target virus, beginning October of that respiratory illness season period (CDC illness season start represented by the black dot-dash line in October). Orange and blue vertical lines with a pointer extend from the dumbbell chart upwards to highlight the timepoint at which target virus was detected on the time-series chart. The colored dashed lines highlight timepoints where a gap in detections between methods was found. Colored solid lines represent timepoints where both methods detected the target virus on the same day. Scatter plots represent the correlation between sequencing-positive rate and qPCR-positive rate taken monthly.

For SARS-CoV-2, the omnipresent nature of the virus in wastewater contributed to its detection by both sequencing and qPCR at the same time across all three seasons **(Fig.6E)**. In addition, no visual seasonal detection pattern was observed for this virus with either technique (Spearman’s ρ = 0.59, p < 0.001). Collectively, this data suggests that over the past 4 years of the program for five different respiratory viruses, the best warning system is a composite of both methods. However, contrary to conventional wisdom that qPCR is more sensitive, sequencing-based methods routinely detected these viruses earlier and more often, likely due to greater sensitivity of capturing diverse nucleic acid fragments across the genome in complex samples.

## Discussion

Molecular detection methods such as qPCR are widely used for viral wastewater surveillance because they provide targeted detection with high sensitivity and specificity. This enables quantification of viral genetic targets, allowing monitoring of community-level virus activity. However, PCR-based approaches require prior knowledge of target genetic sequences, are dependent on sufficient sequence complementarity between target regions and assay primers, and can miss rare, mutated, or other emerging viral pathogens^47,48^. Hybrid-capture sequencing of viral genomes within environmental samples can be used to overcome these pitfalls. Unlike targeted approaches, this method enables detection of multiple viruses (in our program, thousands at one time) and their potential variants while also providing information on viral evolution, all within a single sequencing run. While hybrid-capture sequencing has been increasingly applied to viral wastewater surveillance over the past decade, large-scale, multi-year studies comparing its performance with established targeted molecular assays across thousands of wastewater samples and multiple viruses have not been reported.

To compare targeted- and sequencing-based approaches for WBE, we analyzed qPCR and sequencing outcomes targeting five respiratory viruses; influenza A, influenza B, RSV A, RSV B, and SARS-CoV-2, within nearly 5,000 wastewater samples collected statewide. We evaluated the concordance between methods and independently compared each with statewide clinical cases. Both methods correlated positively with each other and with clinical trends, and we also found that qPCR is more likely to detect a virus when greater genomic coverage is recovered from wastewater. Finally, sequencing detected target viruses as early as or earlier than qPCR, highlighting its potential for early detection of seasonal viruses year-round. Thus, our study showcases multiple key results each contributing to the validity of hybrid-capture sequencing as a suitable alternative for viral WBE.

First, we found that both methods vary in how well they detect respiratory viruses in wastewater. While both methods detected SARS-CoV-2 exceptionally well across tested samples (78% of all tested samples), the remaining 4 viruses had poorer detections across methods (62-81% of tested samples returned negative). This could be due to the seasonality of these respiratory viruses, lower persistence of genetic material in wastewater, or variations in shedding. Samples collected during the respiratory illness off-season may genuinely lack respiratory virus material and consequently, neither method detects them in these samples. This contrasted with SARS-CoV-2, which is present throughout the calendar year, enabling its constant detection. Unlike influenza and RSV which replicate primarily in the respiratory tract, SARS-CoV-2 is capable of replication in the gastrointestinal tract making it more likely to be shed into wastewater^49,50^. In addition, this virus is known to be shed in high amounts in stool compared to influenza and RSV, possibly contributing to its ubiquitous nature in wastewater^51,52^. Together, these key properties could contribute to constant detection of SARS-CoV-2 with both methods. Interestingly, our results highlighted differences between qPCR detections across viruses, with lower detection rates for influenza, intermediate rates from RSV and strongest for SARS-CoV-2. These qPCR results align with studies carried out by others, suggesting similar detection rates between sequencing and PCR-based methods while adding temporal resolution and power for robust, multi-year clinical comparison^40,53^. We also observed that both influenza A and B were more often detected by sequencing than qPCR, possibly due to the multi-target nature of hybrid-capture sequencing, where tiled target probes can bind to fragmented genomes and contribute to positive detection.

Next, when correlating samples tested by both methods for each target virus, we observed that both methods correlated positively with each other **(Fig. 3, Supp Fig. 1)**. Across all viruses, the strength of association varied (Spearman’s ρ 0.1-0.7, all p < 0.01 except Influenza B; p = 0.3), indicating that increases in qPCR measurements correspond to increases in sequencing signal. However, the variation in correlation strength suggests that the agreement between methods is virus dependent. Differences in capture efficiency, sequence diversity, persistence of viral reads, false positive and false negative detections, and relative abundance of viral nucleic acid in wastewater may all contribute to the variation in the strength of association between the two detection methods^54^. Our findings are consistent with previous work demonstrating the utility of hybrid-capture sequencing for viral surveillance over targeted methods^40^. Here, we extend this comparative evaluation of hybrid-capture sequencing and targeted methods across thousands of wastewater samples collected over four years, enabling comparative assessment of method performance over multiple seasons, while assessing temporal consistency in detection outcomes.

Third, to assess per-virus detections with qPCR and sequencing against known clinical cases, we correlated each method’s detections against the statewide clinical ED visit rates per virus **(Fig. 4, Supp Fig. 2)**. qPCR detections correlated positively with ED visit rates for all viruses, though the strength varied considerably; RSV showed the weakest association, followed by influenza and SARS-CoV-2 (Spearman’s ρ = 0.29, 0.58, 0.80 respectively. p < 0.001). In contrast, sequencing detections correlated more strongly with ED visit rates across all three viruses and were more pronounced for RSV (Spearman’s ρ = 0.67, 0.70, 0.91 respectively. All p < 0.001). Our findings align with previous studies where qPCR-based viral detections in wastewater were found to peak during periods when local clinical enrollments in the geographic study region increased^55,56^. In addition, separate studies have found that sequencing-based viral detections correlate with clinical enrollments, aligning with our analysis outcomes^16,57^. By comparing both qPCR and sequencing with clinical data, our work provides insights into each method’s capacity to track community viral activity.

Fourth, we assessed the relationship between sequencing-derived genomic span of mapped target viral reads with the likelihood of qPCR running a positive detection for the same virus **(Fig. 5)**. Across all 5 viruses, we found that every 10% increase in the genomic span was associated with higher odds of qPCR detection. Interestingly, the ability of qPCR to successfully detect a target virus was better when the sequenced reads of that virus were more complete. Most sequencing-positive samples had lower genome span levels and proportionally, qPCR detections among these were low. These results highlight two key points. First, sequencing detected target viruses in samples where genomic span was <20% of the reference genome, a range where qPCR consistently failed to detect the same virus. From a detection standpoint, this is highly valuable in the context of public health, where the earliest signal of a rising virus comes from samples with minimal viral coverage. Second, qPCR detection was strongly associated with the extent of genome recovered by sequencing, indicating that when complete genomes persist, specific regions targeted by qPCR remain intact, in turn allowing them to be captured. Ultimately, our results show that sequencing is a more sensitive approach to viral detection in wastewater, capable of positively detecting target viruses circulating at lower levels. This contrasts with qPCR, whose detection ability is proportional to the genomic span of sequenced reads.

Finally, we looked at hybrid-capture sequencings potential to act as an early detection system for respiratory viruses of concern. Monitoring viruses with established seasonal variations provides a baseline for evaluating the potential of sequencing to look at atypical viral activity such as the emergence of novel variants or viruses with previously unknown prevalence patterns. We found that every October (CDC defined starting month of respiratory illness season), sequencing consistently detected targets earlier than qPCR with leads ranging from 6 to 105 days, or the same time as qPCR depending on the yearly season and virus **(Fig. 6)**. For influenza and RSV, the monthly detection rate with both methods followed a seasonal trend, rising in the fall and declining by the following summer. Interestingly, we also observed periods where samples were consistently positive using sequencing, but absent using qPCR. The implications of this analysis are two-fold. Sequencing can detect trace levels of target virus that remain below the detection threshold of qPCR. Hybrid-capture sequencing probes are designed to capture any fragment of viral genomes and can tolerate at least 10% mismatch, and potentially more with target genome binding without loss of capture efficiency. Hence, positive detections are more likely than with qPCR. However, due to this feature, further deconvolution of sequencing-positive hits is necessary for accurate representation of viral composition within a sample. Still, sequencing based monitoring can act as a proxy for early detection of novel viruses. Second, this analysis highlights a limitation of targeted qPCR in the context of early detection. Successful viral detection depends entirely on a single short region to remain intact through wastewater transit and processing. Targeted assays are additionally vulnerable to sequence drift, as mutations accumulating in primer or probe binding sites can reduce or abolish amplification.

For SARS-CoV-2, its ubiquitous presence makes it a difficult target for an early warning system. This virus is consistently present in wastewater and lacks off-seasonal trends^58^, with detection rates with both methods seldom dropping below 80% throughout the study duration. For this virus, changes in viral abundance may provide a more informative indicator of fluctuations than detection rates alone.

While our work highlights the utility of hybrid-capture sequencing for viral detection in wastewater, several limitations exist. Currently, our results are reported as influenza A dynamics and not by individual subtypes such as avian influenza H5N1, H3N2 and H1N1. Future work includes resolution of influenza A detections to lower taxonomic levels for subtype-level epidemiological analyses. Secondly, our study focuses on statewide viral dynamics, assessing all collected samples together. Analyzing data by collection cities and sites is required to understand community-level viral dynamics. In addition, differences in viral shedding rates, persistence of nucleic acid in wastewater, time of collection, and wastewater flow rate could all affect per-method detection outcomes.

Collectively, our findings support the use of hybrid-capture sequencing as both a complementary and a competitive alternative approach to targeted qPCR for longitudinal monitoring of key respiratory viruses. With high target sensitivity, the ability to detect viral variants, and early warning system capabilities, this method can be used to strengthen wastewater-based surveillance, supporting and benefiting public-health initiatives in the process. Although hybrid-capture sequencing costs approximately twice as much as qPCR in our program per sample, its cost per virus detected is substantially lower. On a per-virus level, sequencing is 10-50x less costly than qPCR because multiple viral targets can be detected in the same reaction, eliminating the need for separate reactions.

As the field of wastewater-based epidemiology increases in popularity, we highlight the benefits of metagenomic sequencing in multi-virus detection while concurrently showcasing effectiveness of qPCR-based viral detection. Ultimately, effective wastewater surveillance requires not only the ability to detect these viruses, but also the capacity to assess their changing abundance over time with high sensitivity, all while remaining cost-effective. By combining the complementary strengths of these methods in wastewater surveillance, health researchers can obtain a more comprehensive image of viral dynamics and strengthen evidence available to inform public health decision-making. As such, this integrated approach can offer a practical foundation for long-term viral monitoring and support public health preparedness for future outbreaks and pandemics worldwide.

## Methods / Online Methods

### Wastewater sample collection, transport and processing

Samples collected across wastewater treatment plants in Texas were preserved, shipped and processed as described by Tisza *et al.* 2023^10^. 100-500 mL samples from over 40 sites across 16 Texas cities were collected and stored in 500 mL prelabeled bottles. To hide the identity of sites where sampling was carried out, collection sites were coded upon request of public health officials.

Decontamination of the surface of collection bottles were carried out with 10% bleach, after which samples were sealed in biohazard bags and shipped inside boxes with ice packs and absorbent pads. All samples were transported as such to the Alkek Center for Metagenomics and Microbiome Research at Baylor College of Medicine, Houston, Texas, where they were stored at 4°C until processing.

### Quantitative PCR of Influenza, RSV and SARS-CoV-2

Real-time reverse transcriptase PCR (RT-PCR) was used to detect RNA respiratory viruses, including RSV (types A and B), influenza viruses (types A and B) and SARS-CoV-2 which were detected using CDC FluSC2 assay (https://www.cdc.gov/flu/php/laboratories/influenza-sars-cov-2-multiplex-assay.html)^10,58,59^. Real-time RT-PCR for influenza and SARS-CoV-2 was performed using 5 µl of eluted RNA and 15 µl of TaqPath 1-step Multiplex Master Mix, (A28523, Applied Biosystems) under the following cycling conditions: 25 °C for 2 minutes, 50 °C for 15 minutes, 95 °C for 2 minutes, and 45 cycles of 95 °C for 3 seconds, then 55 °C for 30 seconds on a 7500 Fast Dx Real-Time PCR Instrument (4406985 Applied Biosystems) with SDS version 1.4 software.

Real-time RT-PCR for RSV A and B was performed with Ag-Path One step-RT-PCR Master Mix (4387391 Applied Biosystems) under the following cycling conditions: 45 °C for 10 minutes, 95 °C for 10 minutes, and 45 cycles of 95 °C for 15 seconds, then 55 °C for 1 minute. Samples were considered positive if Ct values were <45. The real-time RT-PCR included negative extraction control and no template controls. A standard curve with amplicons targeting the primers and probe sequences of each target was used to determine the genomic copy numbers for each target. The details of the primers, probes and amplicon sequences are described in **Supp. Table 3**.

### Wastewater sample processing and hybrid-capture sequencing

Prior to sequencing, 50mL of each wastewater sample was first decanted and centrifuged at 3,374 x g for 10 min to separate the solid and liquid fractional elements. Next, the subsequent supernatant was vacuum-filtered with an ion-based cellulose filter paper. The filter paper, which now contains viruses, was placed into a bead-beating tube along with lysis buffer. The tube was run on a homogenizer for 1 min at 5 m/s, rested for 1 min and then ran again for 1 min at 5 m/s. After the completion of the bead beating step, samples were again centrifuged at 14-17 x 1,000 RPM for 2 min. Post-centrifugation, DNA and RNA elements were extracted using the Qiagen QIAamp VIRAL RNA Mini Kit.

Reverse transcription of RNA to cDNA was carried out with Protoscript II First Strand cDNA Synthesis Kit (New England Biolabs Inc.), NEBNext Ultra II Non-Directional RNA Second Strand Module (New England Biolabs Inc.), and Random Primer 6 (New England Biolabs Inc.) before mixing it with DNA for library construction. Libraries were constructed with Twist Library Preparation EF 2.0 Kit and Twist Universal Adaptor System (TWIST Biosciences) using 25 ng of cDNA and DNA mix. The libraries were pooled with 16 samples, totaling 1,500 ng per pool. Probe hybridization was carried out with Twist Comprehensive Viral Research panel (TWIST Biosciences) at 70°C for 16 hrs. Post-capture pools were further amplified with 12 PCR cycles, and final libraries were sequenced on the Illumina NovaSeq 6000 SP flow cell to generate 2×150 bp paired-end sequence reads. Raw data files were thus produced in the binary base call (BCL) format. The raw BCL files were finally converted to FASTQ format and demultiplexed based on dual-index barcodes using the Illumina bcl2fastq software.

### Wastewater sample inclusion criteria and quality control

Over the 4-year period of constant sample collection from May 2022-July 2026, 4,935 samples were collected. In 2022, only 2-10 sites across 3 cities were sampled originally, slowly expanding over time to over 40 sites across 16 cities by July 2026. To account for gradual increase in sampling sites, per-virus abundance values from sequencing, measured as RPKMF (Calculated as **R**eads **P**er **K**ilobase **M**illion **F**iltered reads), were averaged across all contributing sites in each month. This ensured addition of new sites would not artificially inflate RPKMF values per virus.

All samples within a pool were processed with the same library preparation and hybridization protocol, and no additional batch correction was applied.

### Respiratory virus sequence assignment with EsViritu

Demultiplexed FASTQ files were processed with BBDuk (v38.84) for adapter removal, quality trimming (Q25) and PhiX read filtering. We discarded reads with a minimum average Phred quality score of < 23 and read length < 50bp post-trimming. BBMap was used to remove reads that were mapped to PhiX Illumina spike-in or the human refence genome (GCF_000001405.39). The remaining processed reads were classified using EsViritu (v1.1.6) with default settings enabled. The processed reads were then aligned to the complete Virus Pathogen Database (v3.2.4) with minimap2^60^, and alignments with at least 90% average identity across 90% of read length with CoverM were retained. (https://github.com/wwood/CoverM).

Preliminary detections were those virus genomes which covered a minimum of either 1,000 nt or 50% of the genome/segment length. As dereplicated virus sequences can have highly similar genomic regions, consensus sequences from preliminary detections were extracted with samtools consensus (v1.21)^61^. These consensus sequences were then computed pairwise to each with anicalc and clusters were made from sequences with at least 98% identity with aniclust^62^. The longest sequence in the resulting cluster was taken as the final sequence. The sequenced fastq reads were next aligned to a smaller database with references corresponding to the final sequences using the same parameters with minimap2 and CoverM. Final detections were those virus genomes or read segments with at least 1,000 nt or 50% of the genome length.

At low levels in wastewater, the proportion of reads aligning to virus genomes corresponds to the proportion of viral nucleic acid molecules over the rest of the molecules in the sample. Thus, we used RPKMF, calculated as **R**eads **P**er **K**ilobase of reference genome/**M**illion reads passing **F**iltering, as our abundance metric^10^. This is a relative abundance metric, and not an absolute metric for quantification of specific viral particles in a sample. To transform this metric into an absolute value for direct comparison with qPCR, internal spike-in with SV40 was carried out.

Per sample abundance, along with coverage metrics and taxonomic profiles, was then tabulated and merged into a single table per pool for downstream analysis. From the taxonomic profiles, per virus abundance data for influenza A, influenza B, RSV A, RSV B and SARS-CoV-2 for every sample was extracted.

### Internal spike-in standardization with Simian Virus 40 (SV40)

To convert the relative respiratory virus RPKMF measurements from sequencing to an absolute measurement, we added Simian Virus 40 (SV40) as an exogenous process control to all wastewater samples collected from September 2023. Addition of this control prior to nucleic acid extraction provided an internal standard that underwent the same extraction, library preparation, hybridization and sequencing as the endogenous respiratory virus targets.

SV40 reads per sample were identified and quantified using the EsViritu read classifier software used to distinguish the other viruses^34^. To calculate the absolute abundance of each respiratory virus, we divided the RPKMF of each respiratory virus in a sample with the recorded RPKMF of SV40 in the same sample. This yields a ratio proportional to the absolute abundance of viral targets in the sample, which accounts for the sample-to-sample variation in nucleic acid recovery, library preparation efficiency, and sequencing depth, as these factors affect SV40 and endogenous viruses equally.

We calculated the standardized abundance ratios for samples collected between September 2023 to July 2026, and generated SV40-standardized time-series charts to supplement the time-series charts of the clinical vs. sequencing. Since SV40 was not introduced until September 2023, we did not have standardized data for samples collected from May 2022 to August 2023. Nevertheless, the observed concordance of both metrics during the September 2023 to July 2026 period showcases the reliability of using RPKMF throughout the study period to assess viral prevalence.

### Integration of sequencing data with qPCR data per virus

Once qPCR and sequencing were carried out per sample across all tested samples, the resulting output files were joined based on the sample ID. Each sample ID is unique and represents a wastewater sample collected from a specific site on a specific collection date.

While 4,935 samples were collected over the study period, due to mismatches in timeline and processing, only around ∼4,000 samples were tested with both qPCR and sequencing for the 5 respiratory viruses used in our study. In addition, while wastewater samples were collected from May 2022, qPCR was carried out on samples collected from December 2022 onwards. Thus, samples collected prior to December 2022 were excluded from correlation analyses, even though they were treated with sequencing. Thus, all sampled removed from the study were either tested only by one technique, had incomplete metadata, or were not tested for any of the 5 respiratory viruses of concern and were excluded from the study entirely.

After joining the qPCR and sequencing outputs per respiratory virus, each sample was labelled based on whether that respiratory virus was detected by both qPCR and sequencing, neither qPCR nor sequencing, or detected by either one or the other.

Once the metadata was corrected and compiled across all 5 virus data sheets, both qPCR and sequencing data were tested for normality, after which a correlation analysis between the qPCR and the sequencing was carried out along with clinical case comparison and lead-time analysis.

### Data analysis and visualization

All statistical analysis and graphical visualizations were carried out in Python (v3.12.7) using pandas (v2.2.2), Matplotlib (v3.9.2), SciPy (v1.13.1), NumPy (v1.26.4) and statsmodels (v0.14.6)^63–67^. Custom python scripts for data processing and visualization for each figure are accessible in the public GitHub repository (https://github.com/TAILOR-Lab/qPCR-sequencing-correlation-study). Figure panels with minor visual aesthetic adjustments were compiled and made in Inkscape (v1.3.2). Claude Code (Anthropic; Opus 5.5) was used for annotating plots and improving figure readability. The authors reviewed, edited and approved of all content.

### Assessment of data normality for qPCR and sequencing outcomes

First, to check if the recorded qPCR and sequencing data was normally distributed, we applied a Shapiro-Wilk (α = 0.05) test to each dataset for each of the five respiratory viruses. Only non-zero values were tested (qPCR copies/ml and sequencing RPKMF, untransformed) as non-detections were recorded as zero. Both qPCR (copies/ml) and sequencing (RPKMF) values deviated from normality for all viruses (all p < 0.001; **Supp. Table 1**). Thus, further associations between qPCR and sequencing measurements throughout the study were assessed with Spearman’s rank correlation.

### Correlation analysis between sequencing and qPCR

To correlate qPCR results with sequencing of each respiratory virus, we looked at samples where both methods were tested per virus. Among these samples, a virus may be positive or negative depending on the method used and the presence in the sample. We correlated viral copies per ml from qPCR with RPKMF abundances from sequencing.

qPCR replicates were averaged when both were non-zero copies per ml values. If both replicates were 0, recorded copies per ml was zero. If one replicate was non-zero and the other was zero, the recorded copies per ml for that sample was set to the non-zero value as is.

For sequencing, the RPKMF of each of the 5 respiratory viruses were taken per tested sample and used to correlate with the corresponding qPCR copies per ml count for that sample.

Correlation analysis and plotting of scatter plots was carried out in Python using the data collected from samples which were tested with both qPCR and sequencing. For **Figure 3A**, every qPCR and sequencing datapoint for all 5 viruses were used, including when detection was negative in both or in either technique. To assess the correlation between samples where both qPCR and sequencing detected a non-zero amount of virus, we also extracted a subset of positive-only samples and looked at the correlation between the samples **(Supp. Fig. 1A)**. Violin plots were added to the outer axes of the scatter plots to highlight the distribution of data points across all viruses. The top and right violin plots denote the distribution of all non-zero samples (samples where both qPCR and sequencing were non-zero). The bottom and left violin plots highlight the distribution of sequencing negative-qPCR nonzero, and sequencing nonzero-qPCR negative data points respectively.

In addition to correlating all viruses on the same scatter plot, per-virus scatter plots were generated with all data points as well as positive only data points to assess correlation of qPCR and sequencing techniques at the virus level **(Fig. 3B-F)**. Rain cloud distributions were added beneath the positive only violin plots to highlight the distribution of the data for each of the individual virus plots **(Supp. Fig 1B-F)**.

Spearman’s correlation coefficient values along with the number of samples used per scatter plot for each respiratory virus were tabulated **(Supp. Table 2, Sheet 1)**.

### Comparison of sequencing and qPCR with statewide clinical cases

Clinical data was obtained from the Texas Department of State Health Services (DSHS) website^44^. For our study, we used weekly emergency department visits (ED rate) as our unit to measure clinical incidences of influenza, RSV and SARS-COV-2. Statewide emergency department visit data for these viruses was downloaded from the DSHS respiratory virus dashboard and was correlated separately with sequencing and qPCR results from our study.

Before correlating our data with the clinical ED rate data, we standardized our data to fit the format of the clinical data. The ED rate as obtained was taken weekly, and to match our data, we took the average qPCR (copies/ml) and sequencing (RPKMF) outcomes per week for each comparison. In addition, to correctly correlate our per-virus qPCR and sequencing for influenza A, influenza B, RSV A and RSV B data with the singular clinical influenza and RSV data from the DSHS website, we combined our influenza A and influenza B data, as well as the RSV A and RSV B data respectively into a single dataset.

For the qPCR data, average weekly copies/ml for influenza A, influenza B, RSV A, and RSV B were calculated separately. The corresponding influenza A and influenza B values were then summed to obtain a single weekly influenza measurement, while RSV A and RSV B values were summed to obtain a single weekly RSV measurement. For the qPCR data, if detections were negative (recorded as zero), the values were set to the lowest non-zero value (Limit of Detection) across the sampling duration for that virus with qPCR. Since SARS-COV-2 was already analyzed a single virus, its average weekly copies/ml were used as is without any aggregation.

A similar approach was used to consolidate our sequencing data, where the average weekly RPKMF for influenza and RSV was calculated per virus and then combined to get the overall weekly abundance of the viruses for that week. This approach allowed us to obtain weekly qPCR and weekly sequencing data to complement and correlate with the weekly clinical data obtained.

Once the data was prepared, time-series plots were plotted in Python. Spearman’s rank correlation was calculated between clinical cases and qPCR as well as clinical cases and sequencing, following which coefficients were tabulated **(Supp. Table 2, Sheet 2)**.

### Assessing the relationship between qPCR positivity rate and genomic breadth of sequencing reads

Sequencing reads from each sample were mapped to a database of viral reference genomes using EsViritu (v1.1.6 with database v3.2.4). EsViritu reports the abundances of each detected virus in a sample as reads per kilobase per million filtered reads (RPKMF). From the mapping results, we extracted abundance information of specific viruses of interest: Influenza A, Influenza B, RSV A, RSV B and SARS-CoV-2. In addition to RPKMF, EsViritu also reports genomic depth information, defined as the average number of sequenced reads that align to each nucleotide base in 100 equal-width windows per reference sequence (each segment in the case of influenza). From these per-window depths we estimated genomic span, which is the fraction of the reference genome covered by at least one read, using the Lander-Waterman model assumptions^68^.

The Lander-Waterman model assumes that reads are placed randomly and independently along the reference genome, thus resulting in reads covering a given base following a Poisson distribution. Hence, the probability of a base being covered by at least one read is given by the following formulae.

P(a base has zero reads mapped to it) = e^(-per window genomic depth)

P(a base has at least one read mapped to it) = 1 – e^(-per window genomic depth)

Because the depth varied per window of a single reference, the formula was applied within each window and per window probabilities were calculated. The total genome span per target reference was then calculated using a window-width-weighted mean across all windows. If more than one reference genome for the same target virus was detected in a sample (Ex. Influenza A lineages), the sample-level genome span was calculated as the mean of the per-reference values weighted by each reference’s RPKMF value.

To visualize the relationship between genome span and qPCR detection, the sequencing-positive samples were grouped into ten genome span bins of width 0.1 (0-0.1 to 0.9-1.0). Here, each increment of 0.1 indicates a 10% increase in the sequencing breadth of reads mapped to the reference genome. For example, a genome span of 0.95 would indicate that sequenced reads covered 95% of the reference genome the reads were mapped to. The qPCR positivity rate was calculated as the percentage of sequencing positive samples that were also qPCR positive per bin and plotted as a bar chart with number of samples per bin shown on a secondary axis (log scale).

To assess the relationship between qPCR positivity rate and genome span, we fit a logistic regression curve for each virus with qPCR detection (positive /negative as the outcome and genome span as the continuous predictor). Results were reported as Odds Ratios (ORs) per 0.1 increase in genome span with Wald 95% confidence intervals. As samples were collected repeatedly from the same sites over time, observations may not be fully independent. We repeated the analysis in a sensitivity analysis where we clustered standard errors by site and found that all associations remained significant (p ≤ 0.03; **Supp. Table 4, Sheet 2**).

### Lead-time analysis of sequencing vs. qPCR

To identify whether sequencing or qPCR tested positive for the target virus first, we tracked the earliest date which had a sample that tested positive from October (representing the start of the respiratory illness season according to the CDC) of each year. The number of sequencing-positive and qPCR-positive samples per virus were aggregated monthly, calculating the monthly positivity rate for each method. The monthly positivity rate was defined as the number of sequencing- or qPCR-positive samples divided by the total number of samples collected statewide during that month.

The monthly positivity rate was plotted as a function of sample collection month, and the method which positively detected the target respiratory virus first was highlighted along with the lag between the detection with the other method through a dumbbell plot aligned beneath it. If a lag was present between the detection with both methods, the time gap was highlighted with colored vertically dashed lines. If both methods detected the virus in the same month, colored solid lines were used to indicate no detection lag. Monthly detection rates between the methods were also plotted on a scatter plot and Spearman’s rank correlation was calculated between monthly method-specific positive detections for each virus.

## Funding

This work was supported by S.B. 1780, 87th Legislature, 2021 Reg. Sess. (Texas 2021) (E.B, A.W.M, and J.F.P), NIH/NIAID (grant number U19 AI157981) (A.W.M), Baylor College of Medicine Joseph Melnick Seed (A.W.M), and Alkek Foundation Seed (J.F.P). The funders of the study had no role in study design, data collection, data analysis, data interpretation, or writing of the research article.

## Acknowledgements

J.R.C and A.W.M conceived the study. Methodology was developed by J.R.C, H.P, M.C.R, M.J.T, J.D, J.F.P, V.A, P.A.P and A.W.M. Software was designed and implemented by J.R.C, H.P and M.J.T. Data validation, curation and quality control were performed by J.R.C., H.P and J. C. Formal analysis was conducted by J.R.C, H.P, R.K.P, J.D.C, C.R.M and A.W.M. Investigation was carried out by J.R.C. and H.P. The original draft was written by J.R.C, H.P, and A.W.M. Visualization was performed by J.R.C, H.P and A.W.M. Funding was acquired by E.B. and A.W.M. All authors contributed to manuscript review and editing.

## Data availability

Raw sequencing read data are available in the NCBI BioProject database under accession number PRJNA966185. Clinical emergency department visit rate for influenza, RSV and SARS-CoV-2 is publicly available and was downloaded from the Texas DSHS website (https://texas-respiratory-illness-dashboard-txdshsea.hub.arcgis.com/pages/texas-statewide-emergency-department-visits-for-respiratory-illnesses).

Curated wastewater sample data of qPCR and sequencing results for all 5 respiratory viruses used in this study can be accessed via dedicated GitHub repository (https://github.com/TAILOR-Lab/qPCR-sequencing-correlation-study).

All core analysis code used in this study, including the EsViritu viral mapping pipeline (https://github.com/cmmr/EsViritu), is publicly available. Custom code developed for generation of time-series figures, correlation plots, and correlation analyses are also available in the project GitHub repository (https://github.com/TAILOR-Lab/qPCR-sequencing-correlation-study).

## Supplementary Data

**Supp. Fig. 1:** Correlation of positive-detection qPCR *vs.* Sequencing results across all respiratory viruses

**Supp. Fig. 2:** Correlation of concordant qPCR and sequencing results with clinical cases

**Supp. Fig. 3:** qPCR detection rates *vs.* sequencing genome span across all respiratory viruses.

**Supp. Fig. 4:** Seasonal monthly detection trends of Norovirus with qPCR and sequencing

**Supp. Table 1:** Test for normality of non-zero samples across qPCR and sequencing outcomes for all viruses

**Supp. Table 2:** Spearman’s Correlation coefficient between qPCR, Sequencing and clinical outcomes

**Supp. Table 3:** Sequences and concentration of Influenza A, Influenza B, RSV and SARS-CoV-2 primers and probe standards used in RT-qPCR

**Supp. Table 4:** Logistic regression odds ratio of qPCR positivity with increasing Genome span and 95% confidence intervals

## Notes

### Competing Interest Statement

The authors have declared no competing interest.

### Author Declarations

All clinical data used in this study were openly available at the Texas DSHS website and can be downloaded from the following link. https://texas-respiratory-illness-dashboard-txdshsea.hub.arcgis.com/pages/texas-statewide-emergency-department-visits-for-respiratory-illnesses

