## Supplemental Figures 1-4 for "Four Years of Continuous Weekly Wastewater Analysis Reveals Sequencing as a Robust Alternative to PCR for Respiratory Virome Surveillance"

### Supplementary Figures

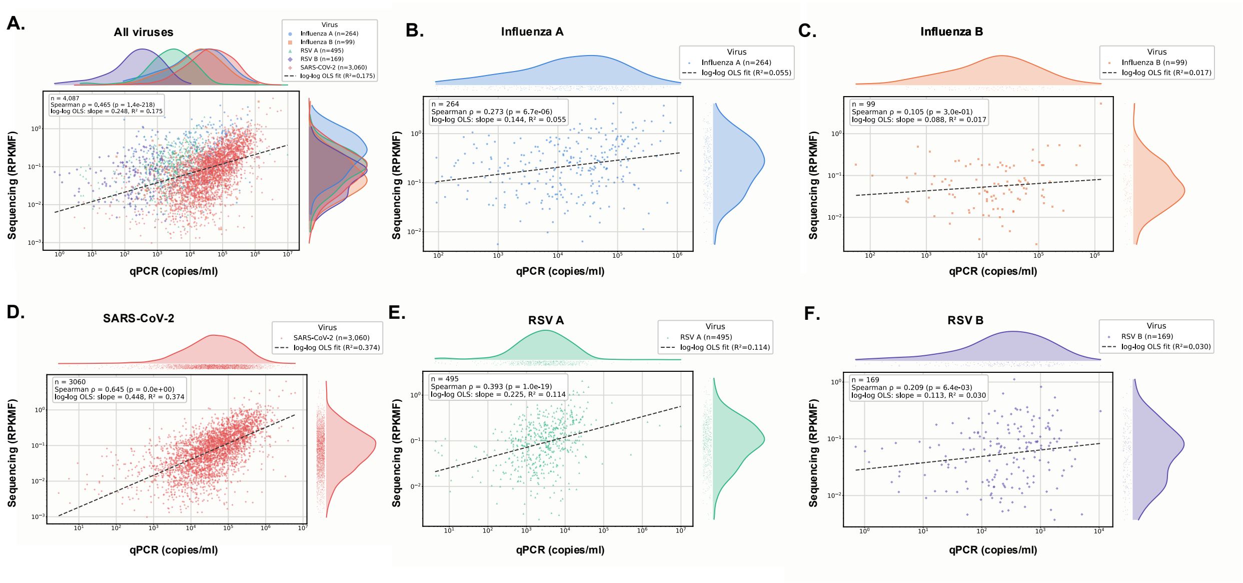

***Supp. Figure 1: Correlation of positive detection qPCR vs. Sequencing results across all respiratory viruses***

*Scatter plots plotted denote the correlation of all samples tested with both qPCR and sequencing for 5 respiratory viruses.* ***(A)*** *denotes the correlation between qPCR and sequencing for all datapoints for all 5 viruses.* ***(B-F)*** *represents per virus correlations for positive-only data points. These include only those samples where both methods detected a non-zero amount of the virus.*

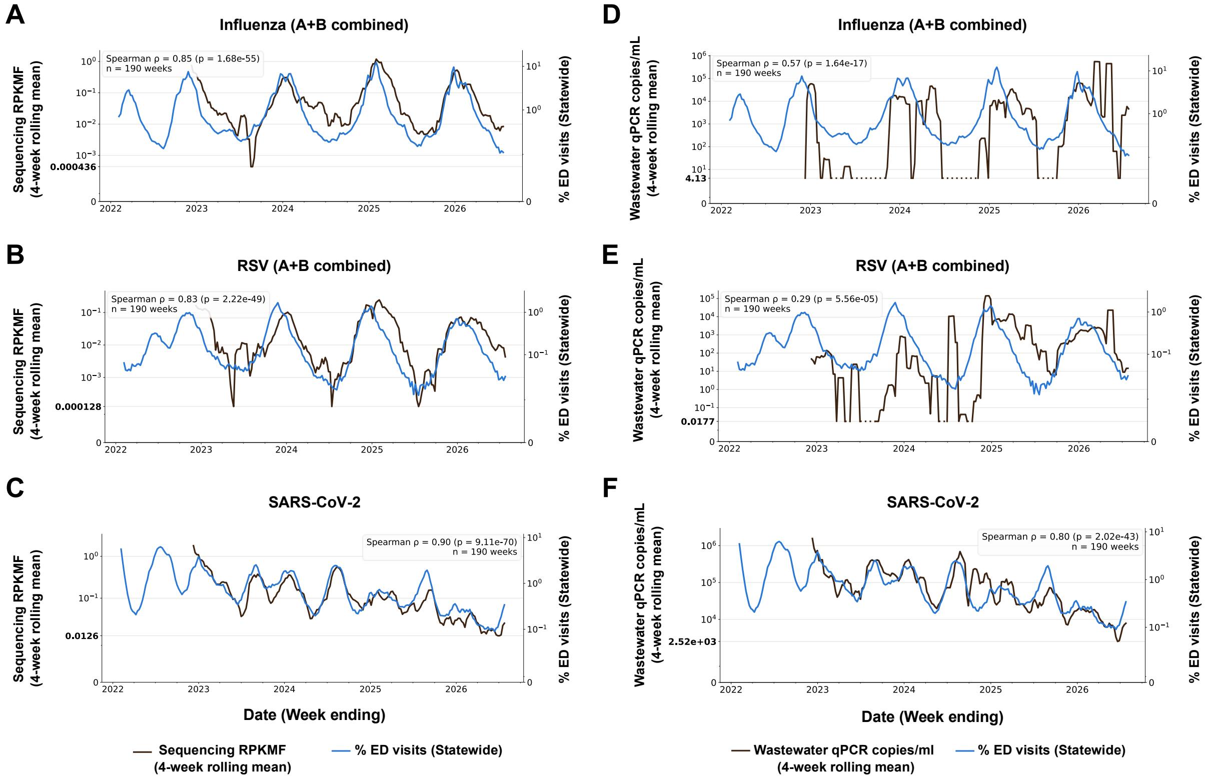


***Supp. Figure 2: Correlation of concordant qPCR and sequencing results with clinical cases***

*Time-series plots represent the correlation of sequencing RPKMF vs. clinical ED rates* ***(A-C)*** *and qPCR copies/ml vs. clinical cases* ***(D-F)*** *for Influenza, RSV and SARS-CoV-2 respectively. All plots showcase only those samples where both qPCR and sequencing were carried out for that virus. The lowest positive detection across the complete time-period is highlighted in bold on the y axis. Negative qPCR detections were set to the lowest non-zero detection.*

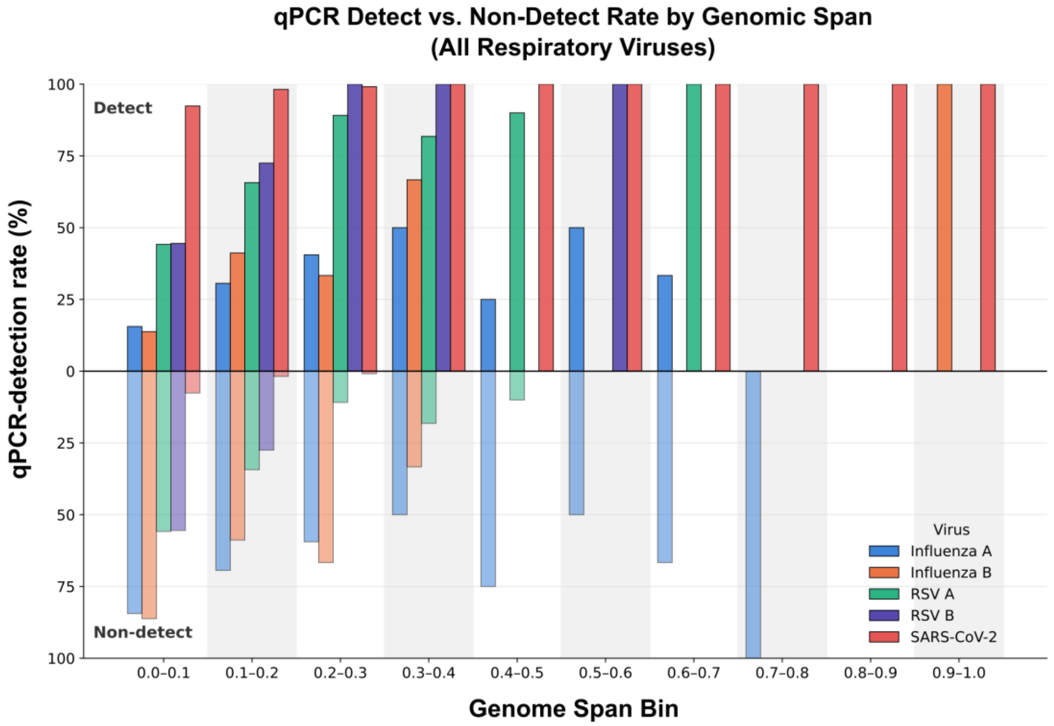


***Supp. Figure 3: qPCR detection rates vs. sequencing genome span across all respiratory viruses.***

*Grouped bar chart represents the proportion of qPCR-positive (above 0) and qPCR-negative (below 0) samples per genome span bin for each respiratory virus. As genome span bin sizes increase, the qPCR positivity rate increases.*

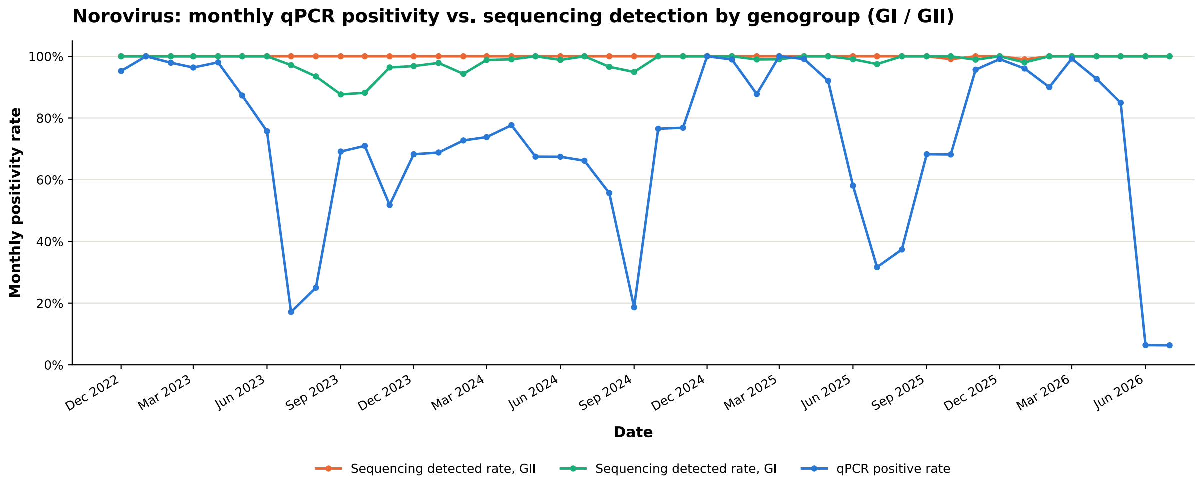


***Supp. Figure 4: Seasonal monthly detection trends of Norovirus with qPCR and sequencing***

*Monthly positivity rate between sequencing and qPCR outcomes for Norovirus were plotted from Dec- 2022 to Jun 2026. Sequencing outcomes for Norovirus were split into GI (Green) and GII (Orange) genogroups to assess variations in sequencing detections. Interestingly, qPCR detections followed a semi-seasonal pattern, while sequencing detections were consistently above 80% throughout the sampling period.*
